# Exploring the KISS principle (Keep It Simple) for decision support systems

**DOI:** 10.1101/2022.08.23.22279126

**Authors:** Anderson Aires Eduardo, Roberto Alves de Sousa, Rafael Maffei Loureiro, Adriano Tachibana, André Pires dos Santos

**Author notes:** Autor correspondente: Anderson Aires Eduardo.

## Abstract

The current existence of massive data has not proved to be sufficient, by itself, for the quality of decision-making in organizations that provide health services. Thus, decision support systems (DSS) have a high strategic potential. However, initiatives focusing on the implementation of such systems commonly frustrate the involved professionals, precisely because of the challenges at data-collection stage. In this context, here we propose a conceptual model of DSS, prioritizing pipelines composed of simple algorithms, presenting low resource consumption for implementation. Our experimental implementation confirmed the computational characteristics preconized by the conceptual model, presenting the potential to mitigate a series of critical points reported by other authors and that negatively impact the real-world implementation of DSSs. Future work should empirically quantify the gains that the implementation of our model can yield, as well as experimentally explore its implementation for more complex organizational scenarios.

## Introduction

Since the advent of internet, back in the 1970s, the evolution of digital technologies has led to a progressive digitization of data and information systems (1,2). Over the last decades, the volume of users (people and organizations) transmitting information digitally has grown at massive rates, at a petabyte scale (2). Currently, the so-called “digital universe” conveys an unprecedented volume of information on a global scale and it is still expanding (1,2).

Naturally, for decision making in any organization, the availability and accessibility of information is a critical factor. This translates into strategic and operational activities closer to an optimal level for each specific situation, enabling greater efficiency and added product value (2,3). However, the current massive flow of data and information along digital systems overwhelms the traditional paradigms of data analysis and processing, bringing new and complex challenges for their application to practical issues (2,4).

In the context of decision-making in healthcare organizations, issues related to obtaining, combining and processing data often have a profound impact on projects to improve the quality of healthcare services (5). On the one hand, successful cases clearly demonstrate the potential of massive use of data, such as the “Google Flu”, capable of predicting flu rates in the USA population by massively analyzing Google search patterns (and achieving a degree of accuracy comparable to the much more complex models of CDC, that country’s center for disease control) (6). On the other hand, there are also numerous cases of failure, often experienced by professionals who work in the routine of organizations that provide healthcare services (5,7). Ironically, the latter case is the most common, especially for health decision support systems (DSS) projects, whose scope encompasses precisely the treatment of large databases to support the quality of decision making (9).

According to Towbin (2019) (5), data collection is the decisive step for the success or the failure of healthcare service-improvement projects. At this stage, critical questions about the nature of the data often go unnoticed by project creators, subsequently impacting their course of development. In the present work, based on the findings of other authors (7,8,9), we propose that this is the main stage of decision support systems implementation projects. Consequently, we also propose that this step should not be cryptic, but instead explicitly allows for the active participation of healthcare professionals. All other steps should be focused on facilitating this one and in the simplest possible way.

In this context, in the present work our aim is to present a conceptual model of DSSs, prioritizing simple, translucid and performative algorithms, while contemplating data collection and database maintenance from the beginning as well as the delivery of recommendations. For the purposes of this work, we experimentally apply this model to radiology prescriptions. Our intention is to counterbalance the natural course of this kind of project, which traditionally involves significant complexity. Following the maxim attributed to Antoine de Saint-Exupéry, “Perfection is not achieved when there is nothing more to add, but when there is nothing left to take away”. The essence of this idea is captured in the principle KISS (acronym for “Keep It Simple, Stupid”), used recurrently in different fields of science and engineering (see (10)). We believe that the proposed model tackles in an integrated way the critical steps for the implementation of DSSs and that it can contribute to the success of projects to improve the quality of health services based on the adoption of these systems.

## Methods

The conceptual model of the proposed decision support system (DSS) is based on an architecture composed of simple and easy-to-implement algorithms. To make this possible, we assume the existence of repositories of professional practice protocols, accessible via the internet. Obtaining the protocol files is performed by a web crawler, actively triggered by the user, whenever the need arises. A second web crawler monitors for updates to the protocol database. We assume that all users have an email account, which can be added to (or removed from) the receiver’s list of updates, to be eventually emailed when modifications are found in the protocols repository. The information from these documents (protocols) is manually tabulated by a team of specialist professionals, forming a database that will be used to parameterize a decision table algorithm (see (11)), which will serve as a knowledge representation model. Recommendations are obtained through user interaction with the decision table algorithm through an interface algorithm, in which basic and clinical patient data are provided by the user (usually, a healthcare professional), who will receive the recommendation of action to be taken. Figure 1 illustrates the proposed conceptual model.

**Figure 1.**
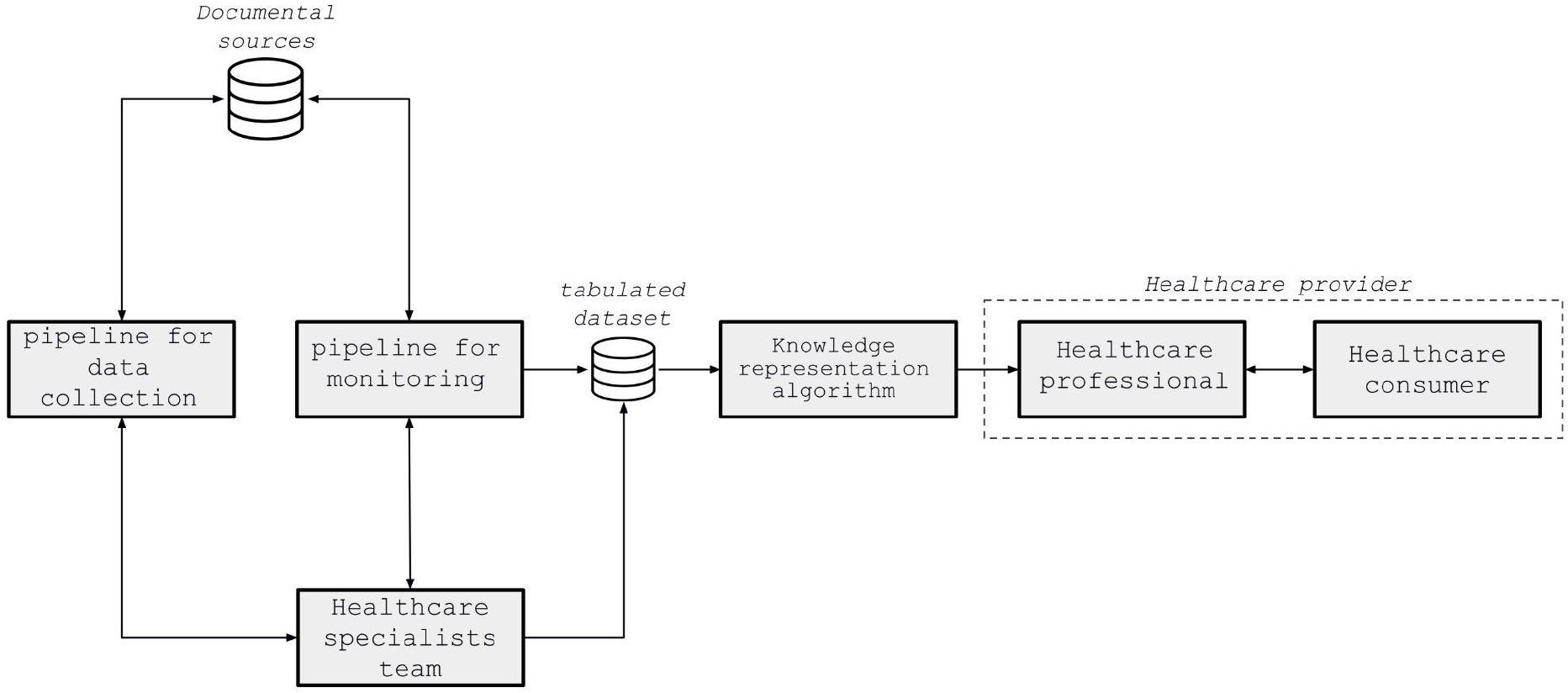
Proposed conceptual model for a decision support system exploring the KISS principle. The data collection and monitoring pipelines are web crawlers. The document sources are repositories of healthcare protocols, accessible via the internet. The team of specialists is an essential part of the model, being formed by healthcare professionals, who work in the focal area of the project to which the system will be implemented.

In the present work, we computationally implemented the proposed conceptual model for the practical case of radiological exams prescription. Thus, the American College of Radiologists (ACR) protocol repository, called ACR Appropriateness Criteria (accessible via the URL https://acsearch.acr.org/list), was used as our document source. The implementation of the web crawlers (`FilesFetcher` and `Monitor`, in Figure 2) and of the decision table (`DecisionTable`, in Figure 02) was carried out as algorithm pipelines using the `python` programming language, version 3.8, using traditional libraries for this purpose. (`beautifulsoup4` version 4.11.1, `pylightxl` version 1.60, plus standard `python` library features). The document’s data tabulation (into the DSS database), for our experimental purposes here, was performed by the authors themselves – being defined head and neck radiology as our focal case. For the interface between system and users, an `API` was implemented, also in `python`, using the `Flask` library (version 1.1.4). two endpoints were implemented, the first being for consultations to the decision table (acepted parameters: *sex, age, clinical indication* and *subcategory* of clinical indication). The second endpoint is dedicated to facilitate the search for *clinical indications* (and their respective *subcategories*) in the system’s database, using an conventional inverted index for that (which was also implemented by ourselves). This `API` was made available through the `AWS Lambda` cloud computing service. For this, we employ the open source `Serverless Framework` tool (see https://www.serverless.com/), which automates the publication of the `API` as a function at `AWS Lambda` and creates a monitoring dashboard. For the purposes of this work, we do not implement any graphical user interfaces, being used the `postman` tool for this purpose (see https://www.postman.com/). Figure 02 illustrates the implemented DSS.

**Figure 2.**
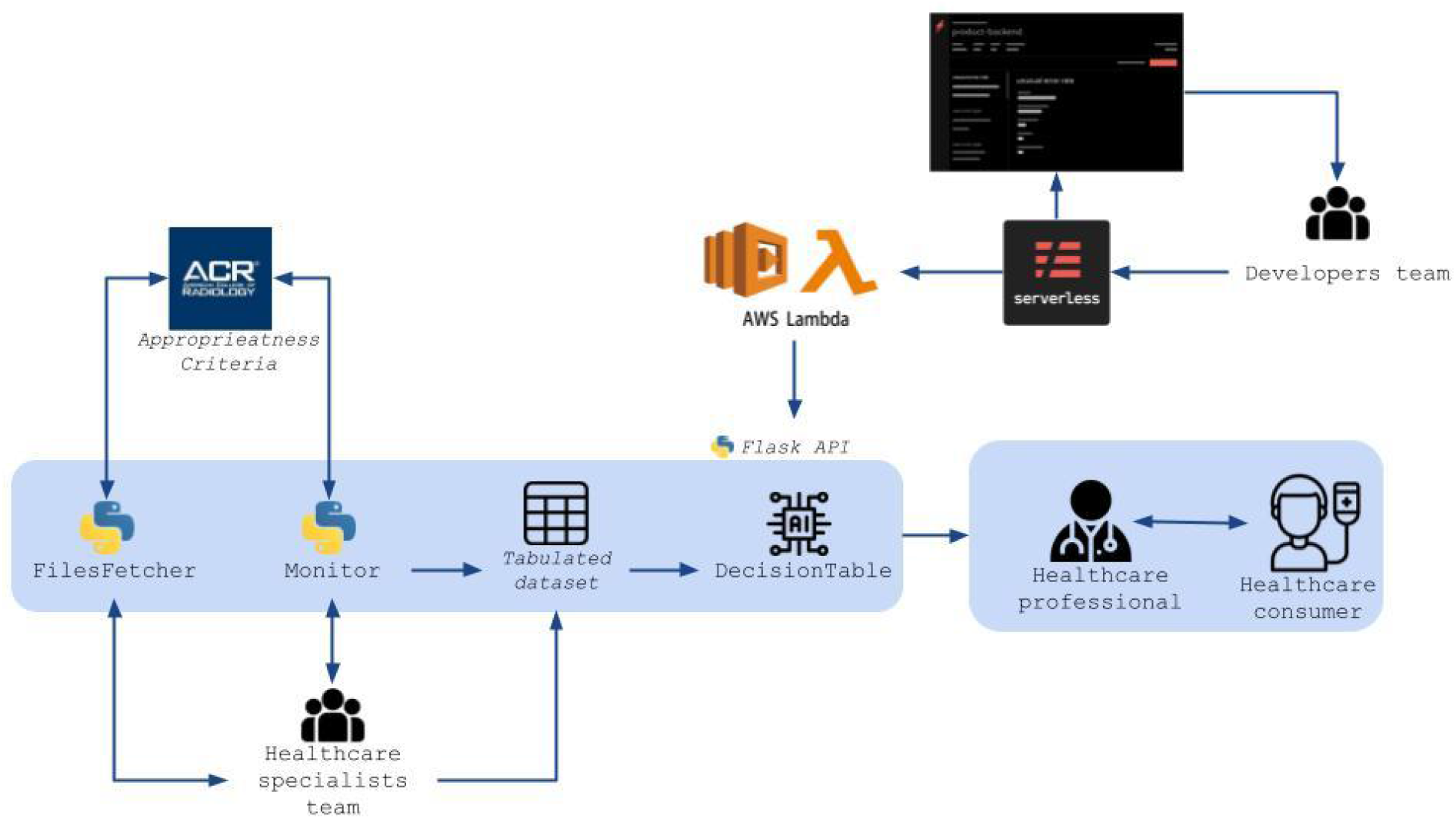
Implemented version of the proposed conceptual model, focusing on head and neck radiology use-case. The role of the team of experts was performed by the authors themselves, for the experimental purposes of this work. The block indicating patient care appears in this figure for illustration purposes only, and was not implemented in our experiment. The entire implementation was carried out using the Information Technology infrastructure of Hospital Israleita Albert Einstein (São Paulo – SP, Brazil).

All steps of this work were carried out using `python` version 3.8, on a personal computer with an `Intel® Core™ i5-10210U` processor, `1.60GHz 2.11GHz CPU` and `16GB RAM`. In view of the replicability of our experimental implementation, the complete source code was made available on the `GitHub` repository https://github.com/AndersonEduardo/kiss-SSD.

## Results and Discussion

The implemented version for the proposed conceptual model proved to be stable and suitable for quick deployment, using the `python` language, combined with the open source `Serverless Framework` tool. The `FilesFetcher` algorithm pipeline proved to be capable of retrieving the files published in the repository used for the experimental implementation (*i*.*e*., the ACR Appropriateness Criteria). A sample of the results is shown in Figure 3.

**Figure 3.**
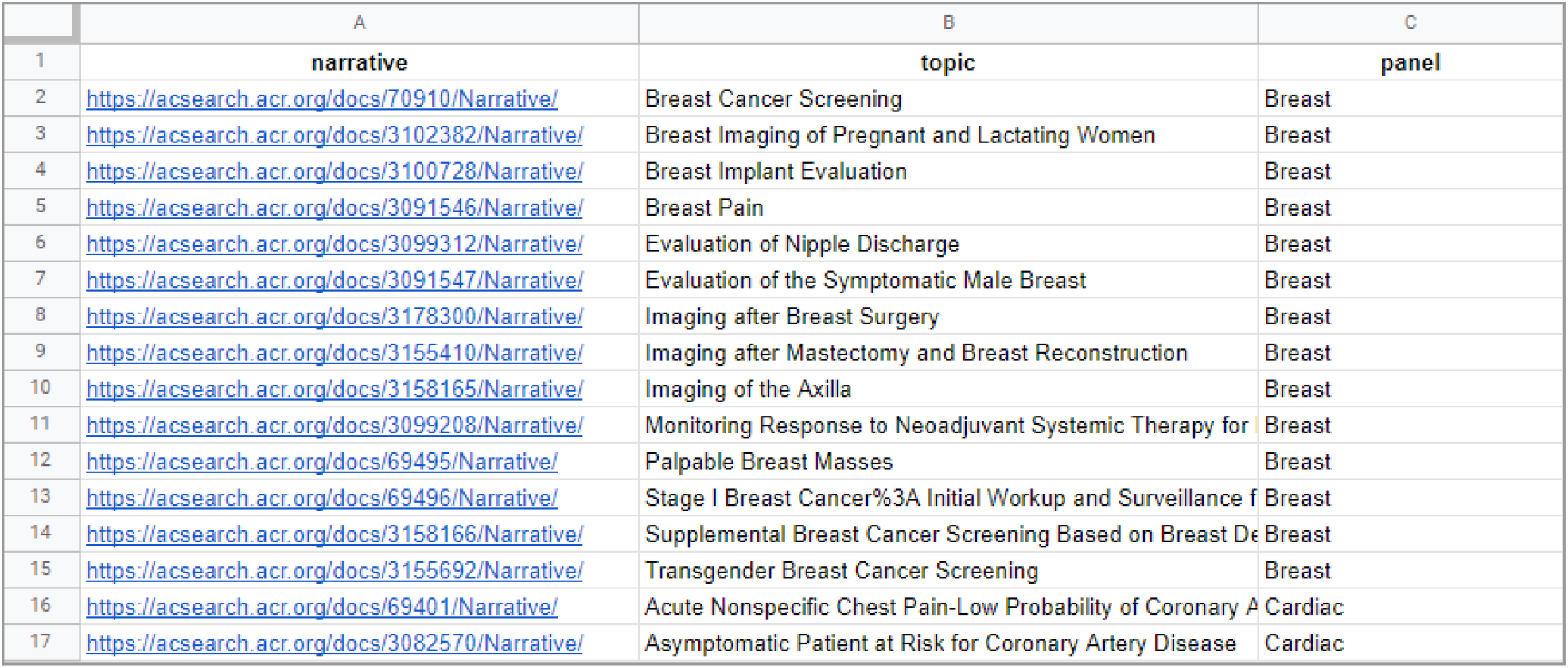
Spreadsheet sample containing the results obtained through the implemented data collection pipeline (`FilesFetcher`). This spreadsheet contains 239 lines and, for convenience, only the first 17 lines are shown (the complete file is available in the project repository - see Methods section).

The `Monitor` pipeline also behaved as expected, performing the checking notifications for updates at the specified frequency (which, for our experimental implementation, was once a week, on Mondays at 6 AM). An illustration of this result can be seen in Figure 4.

**Figure 4.**
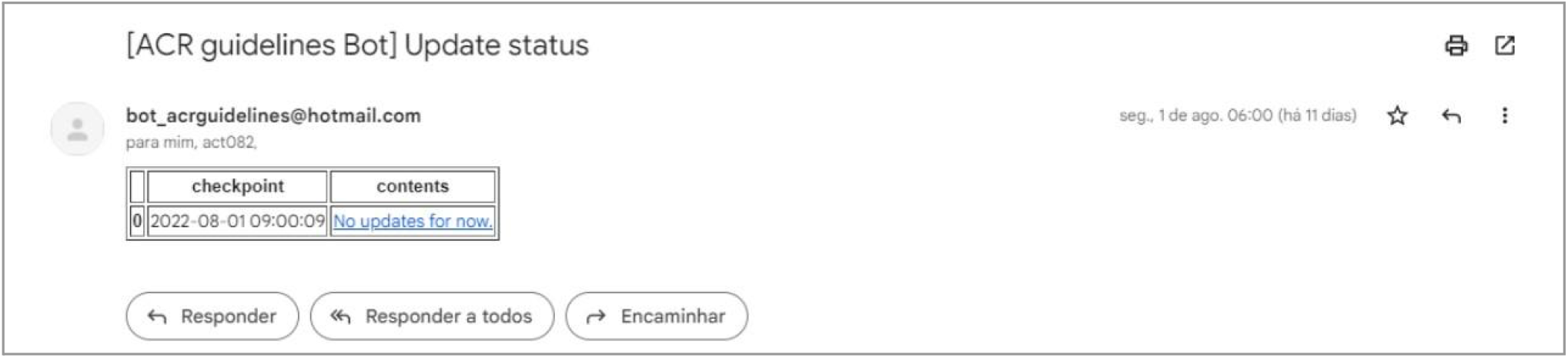
Illustration of the email triggered by the `Monitor` pipeline (for the first author, AAE), which is our implementation for the monitoring module in the conceptual model proposed here. In this image, the email was sent on Monday, August 1, 2022, with no new content found in the monitored radiology protocols web-source. For the implemented version shown here, in situations where protocol updates are found, the document links for each of the updated PDF files are presented in the “contents” column of the table shown in the image.

The implementation of the decision table (`DecisionTable` pipeline), as well as its interface via the `Flask API`, showed results in accordance with the recommendations of the proposed conceptual model. `API` monitoring via the `serverless framework` dashboard also proved to be viable for real-world applications, which often require the deployment of a stable DSS in short periods of time (on a scale of a few days, for example). Figure 5 shows an example of the `API` output and Figure 6 illustrates the `API` monitoring dashboard in action.

**Figure 5.**
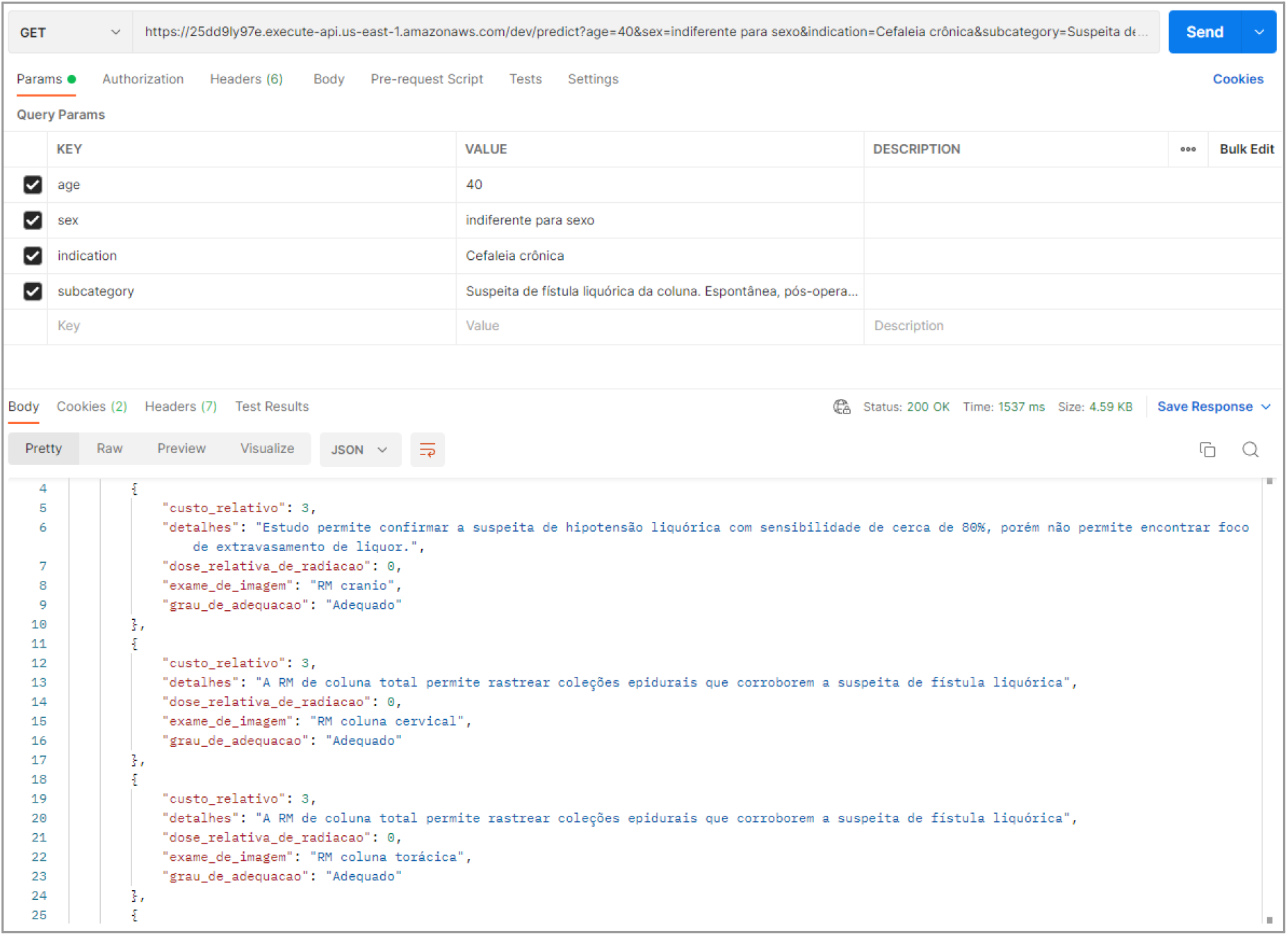
Example of the output for a user query to the DSS implemented upon the proposed conceptual model. The employed parameters were: *age = 40*; *sex = indifferent to sex*; *indication = chronic headache*; *subcatgory = suspected spinal cerebrospinal fluid fistula. Spontaneous, postoperative or post-puncture. Initial image assessment*. For this consultation, 14 indications of radiology exams were returned. For each exam indication, information on the name of the imaging exam, the degree of adequacy, the relative radiation dose, and relative cost, and further exam details are returned.

**Figure 06.**
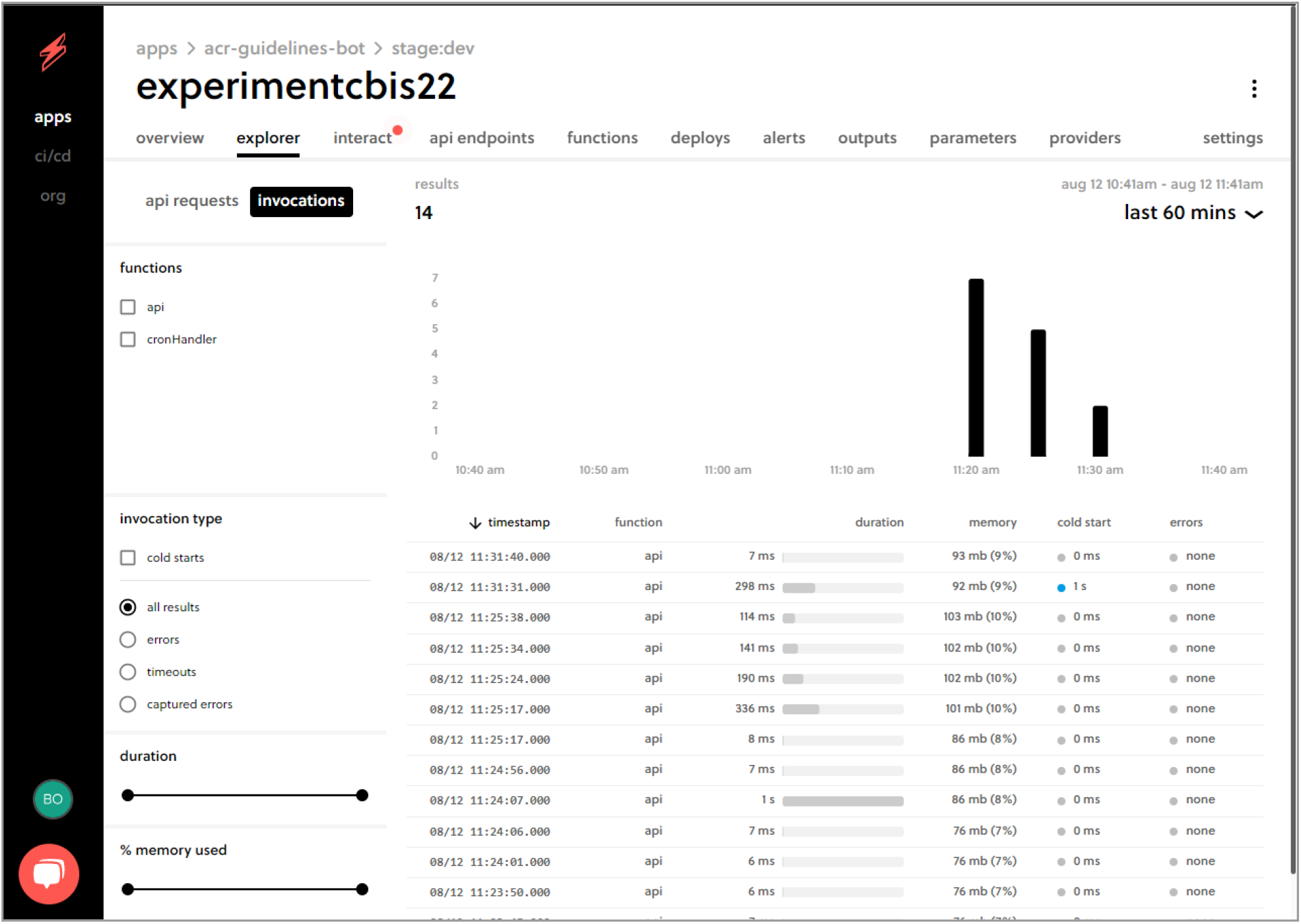
Sample of the `serverless framework` monitoring dashboard for the implemented `API`, based on the proposed conceptual model. From this dashboard, `API` monitoring can be performed by the development team. The image also illustrates the stability of the experimental implementation, showing no errors during the tests performed by the authors.

Taken together, the results obtained allow us to glimpse the potential of the proposed DSS model (exploring the KISS principle) to mitigate critical issues to implement such kind of systems, as identified by other researchers. Kumar (2016)(7) proposes that, in addition to the parts traditionally designed for DSSs (*i*.*e*., hardware, software, data, rules), the “stakeholders” (*i*.*e*., doctors, nurses, laboratory professionals, among others) are also a fundamental part of these systems. Without the participation of these professionals, the implementation of a DSS has considerable potential to lack relevant attributes for the success of its adoption by the focal users. In the model proposed here, the participation of these professionals is a component of the system, when implemented. This feature paves the way to decision support outputs to achieve an ideal situation, in which it is built by health professionals to health professionals themselves.

In addition, as the proposed model prioritizes straightforward algorithms and pipelines, DSS projects based on this model have the potential for low implementation and maintenance costs, maximizing the marginal gains that the implementation of this kind of system can yield to organizations (even in simplified versions). This is an aspect that commonly impacts projects involving the implementation of such systems, being reported as one of the main factors of failure of the initiatives (7,8).

The heterogeneity in the data to be used in the knowledge representation algorithm (here, the decision table algorithm, in the proposed conceptual model) is also a critical point commonly pointed out by different authors (5,7,8) and that can be mitigated in systems that adopt the KISS principle, as the conceptual model proposed here. This potential should be realized when the DSS to be implemented is specific to a particular area or healthcare providing (as in the case of prescribing radiology exams, explored in this paper). This should happen because the system’s database formation relies on the semi-automated compilation of data from documented protocols for professional practice, already validated and agreed by the focal professional community (see the Methods section). Thus, thanks to its modular architecture, the data acquisition pipeline can be customized and implemented to maximally facilitate the things at the manually data-processing stage, considering eventual heterogeneities at document sources. Being the data integration by itself done manually, the aforementioned customization of the protocol acquisition pipeline will not need to deal with all the potential complexity involving the combination of data contents, when coming from different sources. Instead, it should prioritize its delivery to the team of experts within human-readable standards and formats defined in advance and which should ideally be palatable to human manipulation to the maximum extent possible.

Considering our approach here, focusing on a DSS that is both straightforward at implementation stages and meets the practical demands of real-world organizations providing healthcare services, we postulate the existence of limitations, naturally. Firstly, the adoption of our model is limited to contexts in which there is a source of protocols duly documented and accessible via the internet (see the Methods section). Another relevant point is the hybrid nature of the database formation (for more details regarding hybrid databases, see(5)). As already mentioned in this section, this is an aspect that brings benefits (direct involvement of healthcare professionals throughout the implementation of the DSS), but also limits the implementation of highly complex systems, encompassing too heterogeneous, ambiguous and large-volume information contents. In these situations (which are beyond the scope of the model proposed in the present work), more complex systems, specially involving artificial intelligence algorithms, may be more indicated, inevitably (see (8) and (12)).

Future work that seeks to empirically quantify the reduction in the cost of implementing DSSs, when based on the model proposed here, will contribute to the advancement of our research. Furthermore, experiments with the implementation of systems for more complex and larger data sources should also be included in future studies, continuing the results presented here. Finally, the use of the system by a large number of users will also constitute a relevant assessment of the scalability of the DSS model proposed here.

## Conclusions

In this work, we propose a conceptual model for a decision support system, prioritizing straightforward pipelines of algorithms, considering agility at implementation stage and the ability to contribute to the solution of practical problems of organizations that provide healthcare services. Our experimental computational implementation corroborated the aspects advocated by the proposed conceptual model, yielding a stable DSS with the potential to circumvent the main critical issues identified by different authors and that commonly impact the real-world implementation of these systems. Future work should evaluate the potential cost reduction due to the adoption of the proposed model, as well as explore its limitations through experimental implementations for contexts of greater information complexity and user population.

## Data Availability

All data produced are available online at https://github.com/AndersonEduardo/kiss-SSD

https://acsearch.acr.org/list

## References

1. Cerf VG. On the evolution of Internet technologies. Proceedings of the IEEE 2004; 92(9): 1360–1370 doi: https://doi.org/10.1109/JPROC.2004.832974

2. Dash S, Shakyawar SK, Sharma M, Kaushik S. Big data in healthcare: management, analysis and future prospects. Journal of Big Data 2019; 6(1): 1–25 doi: https://doi.org/10.1186/S40537-019-0217-0/FIGURES/6

3. Tajima S, Drugowitsch J, Pouget A. Optimal policy for value-based decision-making. Nature Communications 2016; 7(1): 1–12 doi: https://doi.org/10.1038/ncomms12400

4. Marx V. The big challenges of big data. Nature 2013; 498(7453): 255–260 doi: https://doi.org/10.1038/498255a

5. Towbin AJ. Collecting Data to Facilitate Change. Journal of the American College of Radiology 2019; 16(9): 1248–1253 doi: https://doi.org/10.1016/j.jacr.2019.05.032

6. Ginsberg J, Mohebbi MH, Patel RS, Brammer L, Smolinski MS, Brilliant L. Detecting influenza epidemics using search engine query data. Nature 2009; 457(7232): 1012–1014 doi: https://doi.org/10.1038/nature07634

7. Kumar A. Stakeholder’s Perspective of Clinical Decision Support System. Open Journal of Business and Management 2016; 04(01): 45–50 doi: https://doi.org/10.4236/ojbm.2016.41005

8. Sutton RT, Pincock D, Baumgart DC, Sadowski DC, Fedorak RN, Kroeker KI. An overview of clinical decision support systems: benefits, risks, and strategies for success. NPJ Digital Medicine 2020; 3(1): 17 doi: https://doi.org/10.1038/s41746-020-0221-y

9. Doyle J, Abraham S, Feeney L, Reimer S, Finkelstein A. Clinical decision support for high-cost imaging: A randomized clinical trial. PLOS ONE 2019; 14(3): e0213373 doi: https://doi.org/10.1371/journal.pone.0213373

10. Herzlinger R, Seltzer M, Gaynor M. Applying kiss to healthcare information technology. Computer 2013; 46(11): 72–74 doi: https://doi.org/10.1109/MC.2013.298

11. Eduardo AA, Loureiro RM, Tachibana A, Netto PV, Almeida TF, Monteiro LHAM, Santos AP. Minimal algorithms for knowledge representation in clinical decision support systems research: a theoretical and empirical analysis. medRxiv 2022; 2022.04.20.22274099 doi: https://doi.org/10.1101/2022.04.20.22274099

12. Chen J, Lu C, Huang H, Zhu D, Yang Q, Liu J, Huang Y, Deng A, Han X. Cognitive Computing-Based CDSS in Medical Practice. Health Data Science 2021; 2021:1–13 doi: https://doi.org/10.34133/2021/9819851

